# More than a year after the onset of the CoVid-19 pandemic in the UK: lessons learned from a minimalistic model capturing essential features including social awareness and policy making

**DOI:** 10.1101/2021.04.15.21255510

**Authors:** Miguel A. Durán-Olivencia, Serafim Kalliadasis

**Author notes:** All authors contributed equally to this work. The authors declare no competing interest. E-mails: Miguel A. Durán-Olivencia; Serafim Kalliadasis.

## Abstract

The number of new daily SARS-CoV-2 infections experienced an abrupt increase during the last quarter of 2020 in almost every European country. The phenomenological explanation offered was a new mutation of the virus, first identified in the UK. We use publicly available data in combination with a time-delayed controlled SIR model, which captures the effects of preventive measures and concomitant social response on the spreading of the virus. The model, which has a unique transmission rate, enables us to reproduce the waves of infection occurred in the UK. This suggests that the new SARS-CoV-2 UK variant is as transmissible as previous strains. Our findings reveal that the sudden surge in cases was in fact related to the relaxation of preventive measures and social awareness. We also simulate the combined effects of restrictions and vaccination campaigns in 2021, demonstrating that lockdown policies are not fully effective to flatten the curve; fully effective mitigation can only be achieved via a vigorous vaccination campaign. As a matter of fact, incorporating recent data about vaccine efficacy, our simulations advocate that the UK might have overcome the worse of the CoVid-19 pandemic, provided that the vaccination campaign maintains a rate of approximately 140k jabs per day.

---

The years 2020-2021 have been marked by the extraordinary CoVid-19 pandemic, and the exceptional social measures and restrictions needed to control the spread of the disease. Not surprisingly, a considerable amount of research has been devoted to trying to forecast the evolution of the pandemic with the principal aim of anticipating new waves of infections (1). The most widely-used framework for epidemic model development and computational exploration splits society into three main categories (so-called compartments) according to their status with respect to the disease: susceptible (S), infectious (I) and recovered (R). A natural balance-oriented reasoning, originally put forward in the late 1920s by Kermack and McKendrick (2), can then be used to obtain the so-called SIR model (Fig. 1.a), a nonlinear ordinary differential equation describing the time evolution of the population of the three categories, interconnected via two parameters: the transmission (*β*) and recovery (*α*) rates. The ratio of these two give rise to the famous basic reproductive 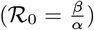 rate. The simplicity and elegance of the SIR have made it a popular generic prototype for numerical and mathematical scrutiny. However, the model also introduces some important limitations which hamper its wide applicability. Specifically, it can only account for a single epidemic outbreak, and only takes into account infection and recovering events. To begin with, the contagious encounters are tacitly assumed to take effect automatically, i.e.whenever a susceptible person meets an infected one, the former becomes infected. At the same time, the original SIR model cannot account for the effects associated with the social preventive response which characterises the *new normal*, e.g. social distancing, mask wearing, limited commuting, remote working, or local curfews and lockdowns, to name but a few examples. These limitations will in turn impact the estimation of the intrinsic parameters *β* and *ℛ*_0_ by using SIR-like models. For instance, different values of *ℛ*_0_ are obtained for the same virus (hence the same inherent properties) under different social contexts, e.g. partial and full lockdown.

**Fig. 1.**
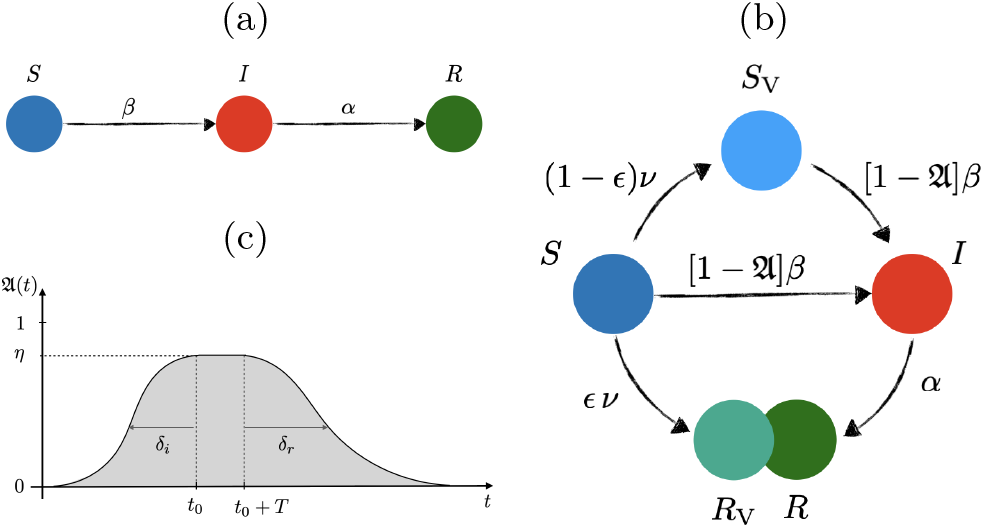
Sketch of transitions in (a) free and (b) controlled SIR network model of disease transmission, and (c) preventive social response, 𝔘(*t*).

Despite the limitations, good fitting can be achieved over limited temporal windows (3–5). However, two serious drawbacks compromise the accuracy of such predictions: a) one cannot fit the whole temporal series, characterised by multiple infection waves, indeed the fit would eventually diverge; and b) the SIR model would never forecast a second or further upsurge in cases. Substantially refined versions of the SIR model have been put forward recently with the aim of including additional important effects, such as shield immunity (6) or exposure to the virus (7). These models are highly intelligent and mathematically elegant, however, their underlying assumption remains that infection occurs automatically. This in turn will necessarily restrict their time domain of applicability. It is clear that more work is needed. In particular, relaxing the assumption of automatic infection raises the exciting possibility of capturing the entire pandemic evolution from day zero up to date and accounting for the multiple waves of infections already experienced. Indeed, to determine whether new variants of the SARS-CoV-2 are more transmissible than their predecessors, the data analysis must cover the entire pandemic outbreak and include the effects of preventive measures and contagion policies adopted by populations and governments, respectively. Many studies also fitted SIR-like models to data from the last stages of the first wave – even though as highlighted earlier the models suffer from the inherent limitation of a single-wave prediction – thus effectively assuming that the epidemic was coming to an end. Yet, it was already known at the time that the number of cases was decreasing because of the preventive measures which in turn should have been sufficient to abandon the corresponding models. Thus, an alternative approach is called for.

The present work is set out as follows. Consideration of the full-history of the data with a *controlled* SIR model (Fig. 1.b) avoids the drawbacks of previous models, by capturing the essence of how the new normal affects the number of infected people. This unveils unique and constant *β* and *ℛ*_0_ for the entire pandemic. Thousands of mutations have emerged in the SARS-CoV-2 genome since the first outbreak in 2019, but the UK strain, known as B.1.1.7, was reported at the time as a more “aggressive” form of the virus, because of an alarming surge in new cases thought to be correlated with the new UK variant, and was one of the main reasons for the lockdown imposed in the UK at the beginning of 2021, e.g. Ref. (8). According to the law of parsimony: chose the simplest explanation from those that fit. Indeed, our results show that the fierce increase in cases is captured without the need of a more transmissible variant. We then put forward the hypothesis that genomic data during the pandemic might have been overinterpretted. To test this hypothesis we carry out a succinct analysis of data from a recent study which links B.1.1.7 with significantly higher viral loads and claims to provide evidence on why transmission was accelerating (9). As far as our particular modelling approach is concerned, its novelty is to include characteristic parameters which could be pivotal in the decision-making process in the coming months. For instance, there seems to be an *inertia of society* which plays a crucial role on the flattening of the curve. For preventive measures to be effective, these should be encouraged quite early in the surge of cases, taking into consideration the inherent social inertia, which typically leads up to a three-weak delay up until society gets to its maximum level of alert. We also account for the effect of vaccination and show that *social relaxation* as of April 2021 without fulfilling a sufficient vaccination rate (determined below) will lead to a new wave of infections over May-June 2021, independently of the more strict lockdown currently imposed since January 2021 and relaxed gradually as of April 12, 2021.

## Minimalistic model including social awareness and policy making

### The model

The controlled SIR model shown in Fig. 1.b (see Methods for mathematical details) incorporates the social awareness and policy making effects on the spreading of the SARS-CoV-2. The main modifications to the basic SIR model are: embodiment of control over the number of infectious interactions that might occur at a given time via the function (1 − 𝔘) which is multiplying the transmission rate *β*, with 𝔘 having the general form showed in Fig. 1.c; inclusion of a time delay for such preventive measures to take effect in reducing the number of effective infectious interactions between infected and susceptible people; accounting for vaccinations not fully effective (ϵ < 1), which in turn leads to the inclusion of two new categories: susceptible after vaccination (*S*_V_), and recovered/removed after vaccination (*R*_V_). These facilitate incorporation of actual vaccination data within the time-evolution simulations of the pandemic, enabling us to estimate the effect that vaccines have had so far, and will have in the next coming months, provided we know the rate of efficacy of the vaccines. Here we take a conservative stance adopting the efficacy value reported for the most widely delivered vaccine so far, i.e. ϵ ∼ 65% (10). As we will illustrate, the minimalistic model we put forward exhibits a distinct predictive capability, and has been rightfully forecasting the evolution of the daily new infections (Fig. 2).

**Fig. 2.**
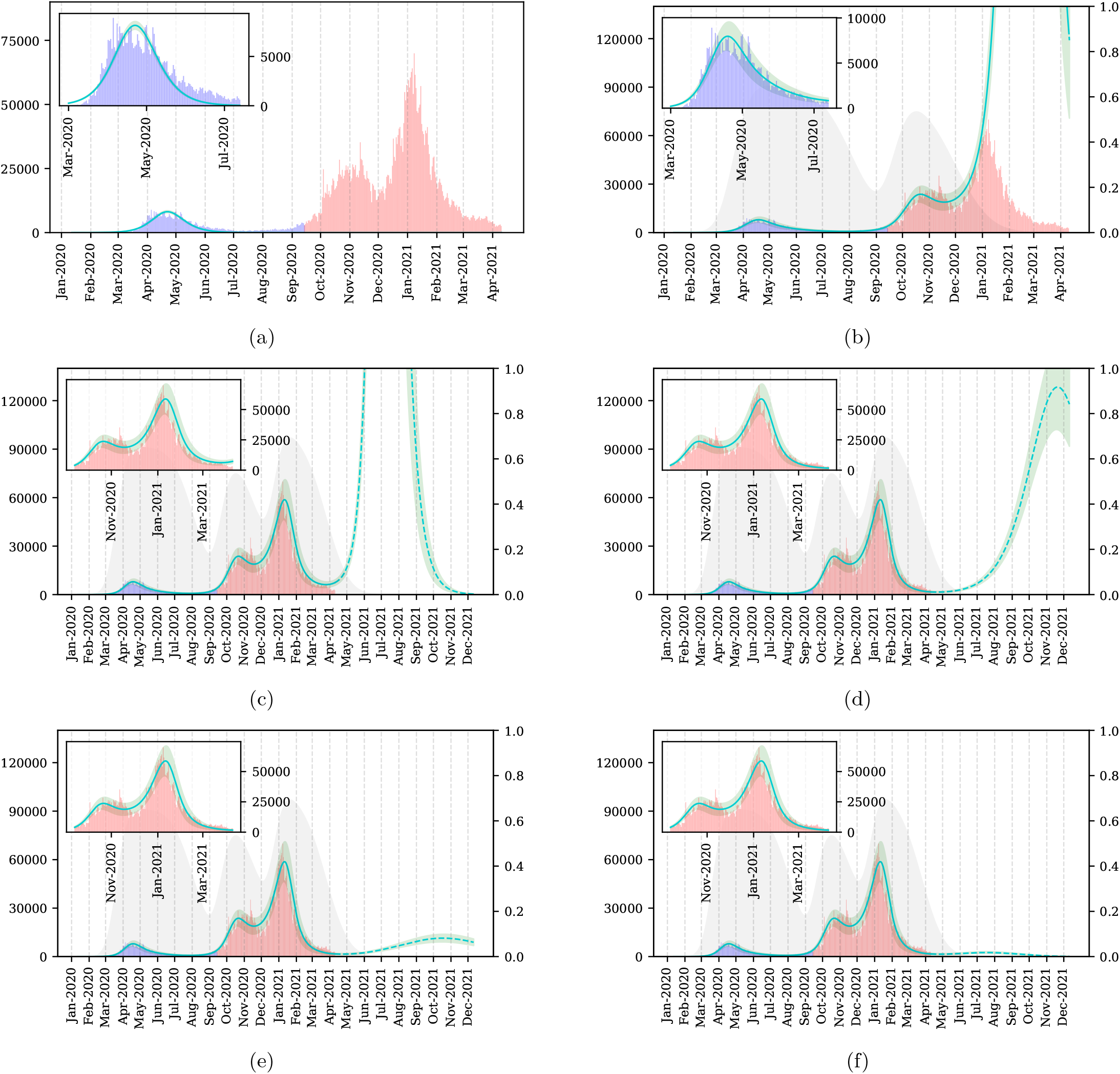
SARS-CoV-2 new daily cases (left axis): blue bars for first-wave data used to fit free and controlled SIR models (light-green lines), red bars for second- and third-wave data used to test the models and their predictions (light-green dashed lines), green areas represent the model uncertainty due to the absolute error of the fitting parameters. (a) Free SIR model captures the essence of the time evolution of new CoVid-19 cases over March-July 2020 (inset plot), but totally fails to predict the second and third waves. (b) Controlled SIR model without vaccination fits better to first-wave data (inset plot) than the free version. The gray area represents the effectiveness of preventive measures, 𝔘(*t*) (right axis). The first wave of social awareness is fit together with *β* and *α*, showing a maximum of effectiveness *η*_1_ ≃ 65%, social inertia *δ*_*i*_ ≃ 21 d, and social relaxation starting at mid June 2020, with prediction of no measures in *δ*_*r*_ ≃ 45 d after relaxation begins. The second wave of social awareness begins in September (confirmed by Prime Minister (12)), reaching *η*_2_ ≃ 60% by mid October 2020 (three-tier system was introduced (13)). The upsurge of CoVid-19 cases in December 2020 is again a consequence of social relaxation. (c)-(e) Controlled SIR model with with a third-wave of preventive measures (reaching maximum effectiveness, *η*_3_= 70%, by the mid January 2021), along with the following casuistry for vaccination: (c) no vaccination, (d)-(f) actual vaccination data from January up to April 10, 2021. Panel (d) shows the effect of stoping vaccination right after April 10, 2021, whilst (e)-(f) reveals a dramatic decrease in infections if maintaining the vaccination rate at ∼ 70 × 10^3^ and ∼ 140 × 10^3^ vaccines per day, respectively. Even with a lower vaccination rate than the mean vaccination rate kept between January and April 2021 (∼ 335 × 10^3^ per day), the UK could be restoring to normal (pre-pandemic) life by June 2021 without risk of a new wave.

### Comparison with observations and predictions

Figure 2 reports curve fits and predictions of the free and controlled SIR models. The free version (Fig. 1.a) fits well the data of the first wave of infections from March to June 2020, but completely fails to predict any second or further wave. This is because in a free SIR model the decay of the infected cases is only possible when the pandemic is already in recession. [As we know now and back in June 2020, this was not the reason for the decrease in cases at the time; rather the reduction of susceptible people was due to preventive measures.] Yet, it has been quite common to use the first wave to extract estimates for *β, α* and *ℛ*_0_. Several works have published estimates for these quantities even by using doubtful methodologies, e.g. in (5), manual fitting of the data to the SIR model was performed, effectively via a trial-and-error approach, on the basis that rigorous non-lineal fitting did not follow the data as well as manual fitting. However, instead of imposing the SIR model and changing the fitting method to achieve agreement with the model, the disagreement with a nonlinear fit is instead a strong indication that the model should have been abandoned.

Evidently, not only does the controlled SIR model (Fig. 1.b) fits better the first-wave data (inset plot, Fig. 2.b), but also captures the underlying reason for the decrease in cases from mid April to August 2020, namely a wave of *social awareness* (𝔘) which effectively reduced the number of susceptible people. The function A embodies both contagion policies and the efforts made by citizens to flatten the curve, e.g. wearing masks, reducing travelling or self-isolating. Moreover, the model correctly predicts a sudden rise in cases when society relaxes, because the downtrend in new infections is not related with the end of the pandemic but with a temporary removal of susceptible candidates from the system. This is precisely what happened from July to September 2020, and what eventually led to the surge in cases in early September 2020. This sharp increase immediately raised the alarm (11, 12), and 𝔘 started growing again, reaching a maximum effectiveness when the three-tier restrictions system was imposed (13). However, these measures were not sufficient to flatten the curve and a new increase appeared in December 2020 because of a gradual relaxation over the month of November. By incorporating new waves of preventive measures in 𝔘, the model is able to reproduce the above observations, as illustrated in Figs 2.b-e.

This provides evidence of the predictive capabilities of the model.

Figures 2.c-d reveal the effects of the third lockdown imposed on January 4, 2021 and of the intensive vaccination campaign followed in the UK (see Fig. 3 for the actual number of doses delivered between January and April 2021). Figure 2.c depicts predictions of what would have happened in case the UK had not delivered any vaccines during the first quarter of 2021. In contrast, Fig. 2.d shows predictions taking into account both the third lockdown just mentioned, and publicly available vaccination campaign data up until April 10, 2021, assuming a conservative 65% efficacy of the vaccine, and stopping vaccination thereafter. As can be seen, had it not been for the very successful vaccination strategy adopted by the UK (exceeding on average the original vaccination target set in January 2021 (14)), the country would be currently facing a new and tremendous wave. This was already forecasted in January 2021 using an early version of the model developed here (15), which was recently confirmed in Ref. (16). In a very recent news article (17) the UK Prime Minister attributed the reduction of new CoVid-19 infections mainly to the the lockdown. While the lockdown has certainly contributed to a temporal slowdown of the curve having an immediate effect, as illustrated in Fig 2.c, it appears that the sustained slowdown of the curve is due to a combined effect of lockdown and vaccination campaign, Figs 2.d-e, with eventual flattening of the curve for sufficiently high vaccination rate, as shown in Fig. 2.f. In fact, Figs 2.e-f uncover a more optimistic scenario for the rest of the year 2021 by simply maintaining a vaccination rate in the range between 70 × 10^3^ d^−1^ and 140 × 10^3^ d^−1^, respectively. What is more, if a target of 140 × 10^3^ d^−1^ is met, the UK could be returning to pre-CoVid-19 normality as early as June 2021.

**Fig. 3.**
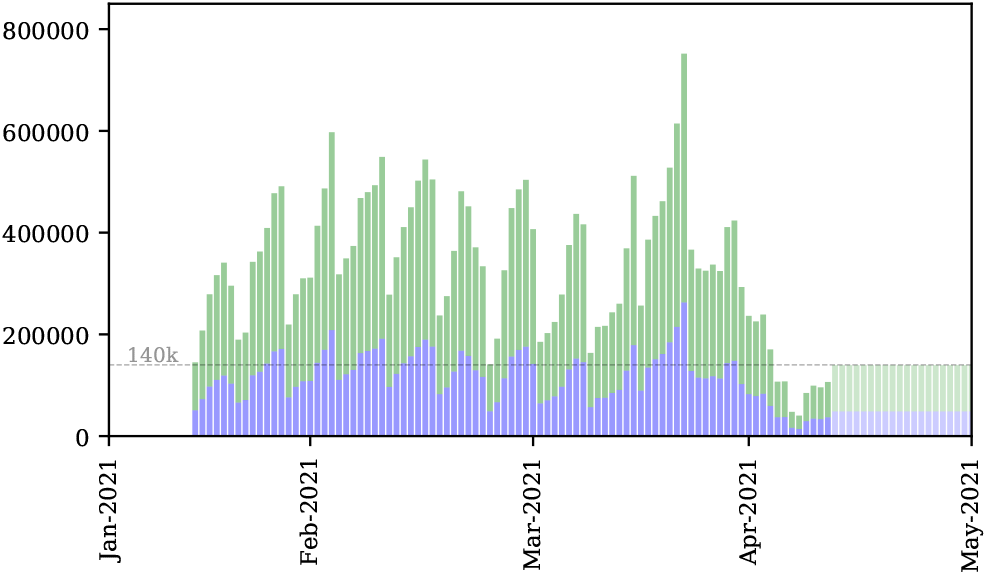
Number of people who have received a first dose CoVid-19 vaccination per day. Split into effective (green) and ineffective (blue) cases has been artificially carried out by utilising an efficacy estimation of ϵ ∼ 65%. The dashed-grey line marks the target vaccination rate of 0.2%d^−1^ (∼ 140k doses per day), also represented as light blue and green bars between April and May 2021 to highlight that these are not real data, as opposed to data from January up to April 2021, taken from official sources.

### Future refinements of the model

There is a number of interesting questions related to the analysis presented here and a discussion of extensions of the model is in order. A first question would be the effect of immunity loss after vaccination, which would involve a new transition *R*_V_ → *S* in Fig. 1.c. This enhancement of the model should not entail a major difficulty and would allow for the study of when and how the pandemic-endemic transition will occur. However, it must wait up until conclusive data is gathered regarding such immunity loss process. Another exciting avenue would be the coupling of the model with a model for the population dynamics, which would permit interrogation of the effects of spatial heterogeneities in the evolution of the pandemic. A similar connection with the basic SIR model has been recently introduced in Ref. (18) by using a generalised diffusion equation, also referred to as dynamical-density functional theory DDFT. This work could be nicely extended by coupling the basic DDFT equation with our controlled SIR model. Additionally, more general DDFT versions can be used to incorporate the effect of mobility anisotropies and “hydrodynamic interactions” (19, 20). Last but not least, the effects of fluctuations, inherent to complex systems, could be accounted for by using fluctuating DDFT as a model for the population dynamics (21, 22).

## Higher-transmission evidence?

### Insufficient statistics in QPCR tests

In a recent study Kidd *et al*. (9) examined the presence of SARS-CoV-2 in respiratory samples using the Thermofisher TaqPath RT-QPCR test. They found that a considerable number of such samples exhibited a characteristic mutation of the lineage B1.1.7, namely the Δ69/70 deletion. This mutation induces a failure in detecting the S-gene (“S-gene negative”) of the TaqPath test, but with two other gene targets clearly detectable: ORF and N. Analysing the RT-qPCR threshold-crossing (*C*_*T*_) values originated from both S-gene negative and positive samples, they obtained the data shown in Fig. 4 for the *C*_*T*_ -value histograms corresponding to ORF-positive samples (in their study both ORF and N-gene targets are shown, but they exhibit similar behavior). Such histograms, where S-negative (B1.1.7) is red and S-positive is blue, were then used to extract the corresponding median *C*_*T*_-values, also shown in Fig. 4: 18.2 (red dashed line) and 22.3 (blue dashed line). From an ensemble point of view, a low *C*_*T*_ value indicates a high concentration of viral genetic material, and vice versa. And it is precisely tis concept the authors used to justify that given the lower median *C*_*T*_ value of the S-negative samples, the UK variant had a higher viral load. Had the S-negative histogram been at least as good and clear as the S-positive histogram, the conclusion that the B1.1.7 samples have a higher viral load could have been defended (whether this means higher transmission rates or not would require further analysis). However, it is clear that the statistics of the S-negative is poor to enable concrete conclusions from the comparison of its median with the S-positive counterpart. Even more so, there is a subtler and more problematic issue when comtrasting *C*_*T*_ values: the lack of infection synchronisation between patients makes it difficult, if not impossible, to unequivocally justify that lower *C*_*T*_ values mean higher transmission rates. For this to be true, the samples should come from patients who have been infected for the same amount of time. Otherwise, by pure chance we could be comparing people at an early stage, of whom a lower viral load, in general, is expected, against patients who have been infected for a longer time, hence with higher viral loads. In any case, it is clear from Fig. 4 that the median *C*_*T*_ values extracted from such histograms are not sufficient to establish a clear difference between the viral-load distributions observed.

**Fig. 4.**
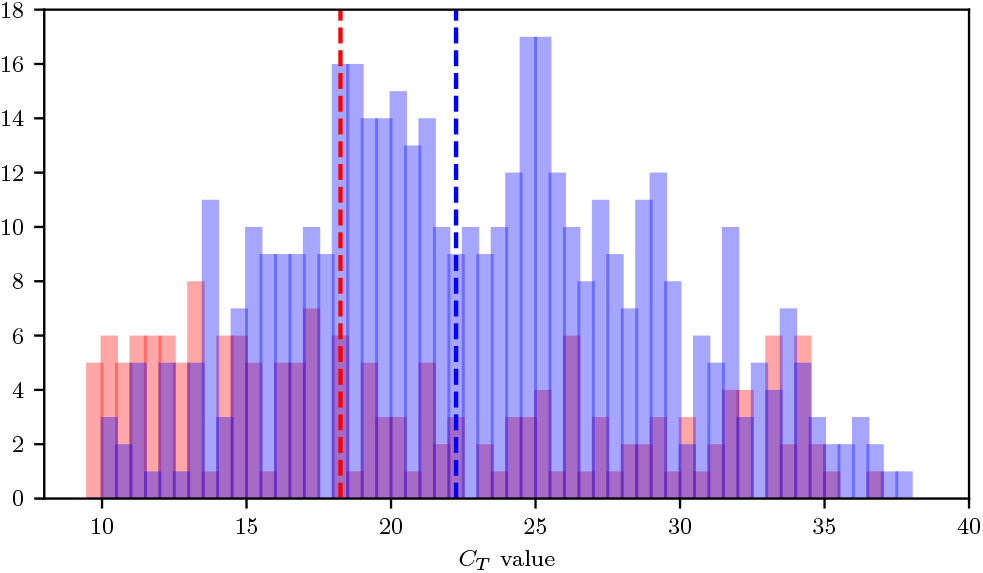
Histogram of *C*_*T*_ values corresponding to S-negative (red bars) and S-positive (blue bars) of ORF positive samples. The median *C*_*T*_ values for each distribution are shown as dashed lines. Data from Ref. (9).

### Enhanced binding does not equal enhanced infection

Protein-protein interactions play a pivotal role in the docking and entry stages of viral infections. In the particular case of SARS-CoV-2, the spike-like protein on the surface of the virus binds to the human cells primarily via interaction with the angiotensin-converting enzyme 2, or simply ACE2 “receptor.” Therefore, ACE2 acts as a cellular doorway for the SARS-CoV-2. Mutations on the spike protein, as is the case of the B1.1.7, can affect the stability of the binding, and hence the infective capability of the virus. Using a large number of molecular dynamics (MD) trajectories of different spike protein interactions with the human target protein (ACE2), a recent in-silico study has reported that mutations of the UK variant has greatly stabilised the interaction with the ACE2, which in turned has enhanced the binding stability (23). The study conjectures that the enhanced binding associated with the spike mutations of the B1.1.7 linage might be responsible for a higher transmissibility of the SARS-CoV-2 virus. However, the binding of the viral particle with the surface receptor of the host cell is only one of the several and complex processes involved in a viral infection (24). For instance, it can happen that the mutation provides the virus with a more stable binding, but following binding the virion remains attached to the cell surface without being internalised (which leads to viral replication) by the cell for longer times, which could potentially result in even slower infection rates. Even if the binding is better and, say, the internalisation effectiveness is the same for all the mutations, this only ensures evolutionary displacement of the lesser stable by the more stable binding, and not higher infection rates. The only way to assert that a given mutation yields a higher infection rate would be by measuring a higher number of virions newly formed inside the cell over a similar time period, which is entirely dependent of the cell machinery itself. Thus, by only having evidence of an improved binding capacity, it seems reasonable to assume that the same amount of virions per second will be produced by using the same cell machinery, which means the same infection rate. In fact, the improved binding associated with the UK variant offers a good justification for the dominance of the B1.1.7 linage observed elsewhere (25, 26), but it does not represent a good basis to justify a higher transmissibility/infection rate.

## Methods

### Population dynamics

The population is split into four groups: susceptible (*S*), infected (*I*), recovered (*R*) and vaccinated (*V*), as illustrated in Fig.1. The vaccinated group consists of two sub-groups: susceptible (*S*_V_) due to ineffective vaccination; and recovered (*R*_V_) due to effective vaccination. The groups follow the delayed dynamical system:

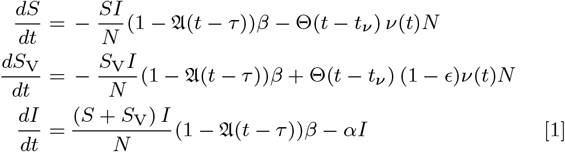

where 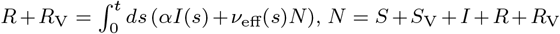 is the total population, assumed *N* ≈ 66.6 million, Θ(*t* − *t*_*ν*_) is the Heaviside step function, *t*_*ν*_is the onset of the vaccination campaign, *ν* is the vaccination rate per day normalised to the total population, and *ν*_eff_ = *ϵν*(*t*) with *ϵ* ∈ [0, 1] is the vaccine efficacy. The parameters *β* and *α*, are the transmission and recovery rates, respectively, and the function 𝔘 (*t* −*τ*) is the percentage of susceptible people using effective preventive measures at time *t* − *τ* (*τ* being a characteristic time for such preventive measures to become apparent, assumed 14 days, the incubation period (27)), with general functional form: 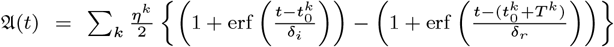, where *η*^*k*^ ∈ [0, 1] is the effectiveness of the preventive measures taken in the *k* − th wave, *T*^*k*^ is related to the time extension of these measures, and *δ*_*i,r*_ are the social inertia (*i*) and relaxation (*r*) time scales, respectively. With 𝔘 = 0 and *ν* = 0, Eq. (1) becomes the free SIR model. For 𝔘 ≠ 0 we get a controlled SIR model. The initial condition used: *I* _0_ = 1 (number of infective cases reported on January 11, 2020), *S*_0_ = *N* − *I*_0_ and *R*_0_ = *V*_0_ = 0. Thus, fitting of five parameters is needed, and this is done at the first wave only.

### Training and testing the model

The full dataset, *Y*, was renormalised to 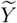, by using a linear fit, *z* = *b* + *mx*, to the number of daily CoVid-19 tests per thousand people given in Ref. (28), with *b* = 0 and *m* = 0.0191 test*/*10^3^people*/*d (from 0 to 7 test*/*10^3^people in 366 days), so that 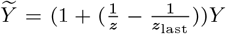. We then use non-linear least squares to fit the daily new cases 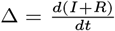 to the first-wave data (training dataset). This fitting yields for the free model: *β* = 0.475d^*−*1^ and *α* = 0.381d^*−*1^ so that 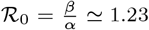. For the controlled model we get: *β* = 0.209 d^*−*1^, *α* = 0.102 d^*−*1^, *η* = 0.65, *δ*_*i*_ = 21 d and *δ*_*r*_ = 45 d, which yields *ℛ* _0_ ≃ 2.042. To test the models we numerically integrate Eq. (1) for both cases, i.e., 𝔘(*t*) = 0 and 𝔘(*t*) ≠ 0. The controlled-model prediction for new infections grows exponentially as of September 2020 (testing dataset) when the first wave of preventive measures would vanish according to the summer trend. With *δ*_*i,r*_ fixed from the first wave, we fit the parameters *η* and *T* of a second social response to unveil the behavioural changes adopted against the apparent second wave of cases, obtaining a maximum of social response by mid-end October 2020. This appears to be in agreement with the declaration of the UK Prime Minister of “seeing a second wave” on September 18, 2020 (12), and his statement on coronavirus where the three-tier restrictions system was imposed, October 12, 2020 (13). For predictions as of January 2021, we introduce a third wave of measures with *η* = 0.70, starting in January and ending in April 2021, *T* = 90 d, which represents the current contagion policies being taken by the UK government. Finally, we use the official number of first doses delivered in the UK in conjuction with two scenarios as of April 10, 2021: 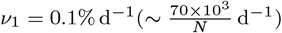 and 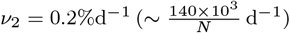.

### Error propagation and uncertainty quantification

For the computation of the uncertainty areas showed along with the model simulations in Fig. 2, we used the common first-order truncation of the Taylor’s expansion of the law of propagation of uncertainty method (29, 30), given the numerical errors calculated after fitting the parameters of the model to data during the training stage.

## Data Availability

All data for the analysis was collected from
https://coronavirus.data.gov.uk/. Specifically,
new cases: https://coronavirus.data.gov.uk/details/cases, and vaccinations: https://coronavirus.data.gov.uk/details/vaccinations.

https://coronavirus.data.gov.uk/details/cases

## Data Availability

All data for the analysis was collected from https://coronavirus.data.gov.uk/. Specifically, new cases: https://coronavirus.data.gov.uk/details/cases, and vaccinations: https://coronavirus.data.gov.uk/details/vaccinations.

## Code Availability

The code used in the creation of this manuscript is available at https://github.com/migduroli/covid-uk-variant/. Model simulations were numerically integrated using odeint from scipy (31), a Python wrapper for LSODA from the FORTRAN library ODEPACK (32). The non-linear square fittings where carried out by using curve_fit from scipy (31).

## ACKNOWLEDGMENTS

We acknowledge financial support from the Engineering and Physical Sciences Research Council of the UK via Platform Grant No. EP/L020564/1.

